# Evidence for protein leverage on Total Energy Intake, but not Body Mass Index, in a large cohort of older adults

**DOI:** 10.1101/2023.06.14.23291391

**Authors:** Sèwanou H. Honfo, Alistair M. Senior, Véronique Legault, Nancy Presse, Valérie Turcot, Pierrette Gaudreau, Stephen J. Simpson, David Raubenheimer, Alan A. Cohen

**Author notes:** COMPETING INTERESTS The authors declared no conflict of interest. CONTACT INFO: PRIMUS Research Group, Department of Family Medicine, University of Sherbrooke, 3001 12e Ave N, Sherbrooke, QC J1H 5N4.

## Abstract

**BACKGROUND:** Protein leverage (PL), the phenomenon of food consuming until absolute intake of protein meets a target value, regardless of shortfall or overconsuming for other nutrients in the diet and total energy intake (TEI). Evidence for PL was observed in humans, recently in a cohort of youth with obesity. This study aimed to test for PL and the protein leverage hypothesis (PLH) in a cohort of older adults.

**METHODS:** We conducted a retrospective analysis of dietary intake in a cohort of 1699 community-dwelling older adults aged 67-84 years from the NuAge cohort. We computed TEI and the energy contribution (EC) from each macronutrient. The strength of leverage of macronutrients was assessed through power functions (*TEI* = *µ * EC^L^*). Body mass index (BMI) was calculated, and mixture models were fitted to predict TEI and BMI from macronutrient ECs.

**RESULTS:** The mean TEI was 7,673 kJ and macronutrient ECs were 50.4 %, 33.2 % and 16.4 %, respectively for carbohydrates, fat, and protein. High carbohydrate intake was associated with low fat intake. There was a strong negative association (*L* = -0.37; p < 0.001) between the protein EC and TEI. Each percent of energy intake from protein reduced TEI by 77 kJ on average, *ceteris paribus*. BMI was unassociated with TEI in this cohort, so the PLH could not be tested here.

**CONCLUSIONS:** Findings indicate clear evidence for PL on TEI, but not on BMI, likely because TEI and BMI become increasingly uncoupled during aging.

## INTRODUCTION

The macronutrients – carbohydrates, protein, and fat – are the main sources of daily energy intake [1–2] in the general population, augmented by energy that may come from alcohol and soluble fibres. Consequently, the regulation of macronutrient intakes and their relationship with total energy intake (TEI) is central to metabolic science. For each gram catabolised, carbohydrates and proteins provide 16.7 kJ (4 kcal), while fats yield around 37.7 kJ (9 kcal) [3]. Given that long term calorie intake in excess of calorie expenditure can cause overweight/obesity to develop [4] and metabolic/inflammatory complications [5], understanding the way macronutrient intake can intrinsically regulate energy intake would help optimize nutritional counselling.

*A priori* there is a daily energy intake that needs to be achieved for the proper functioning of an organism, which depends on factors such as physiological state, health, and morphology [4]. However, beyond supplying basal energy, the different macronutrients also have their own independent and interactive effects on health and appetite. The geometric framework for nutrition is an approach that was developed to help disentangle the relevant relationships between nutrients, food environments, the composition of foods and physiology. The geometric framework for nutrition uses an *n*-dimensional state-space where each dimension represents a food component of interest [6, 4, 7]. In practice, two or three nutritional dimensions usually related to macronutrients are considered.

Geometric framework for nutrition experiments involving intake measured on foods that vary systematically in the composition of *n*-nutrients of interest have demonstrated that animals do not feed on macronutrients indiscriminately. Rather, different organisms display specific macronutrient appetites and “intake targets” [8]. When consuming foods that are imbalanced relative to the nutrient ratio of a target, intake must be compromised based on rules that may lead to excesses of some nutrients and deficits of others. A range of different rules have been observed in different species; however, several organisms display ‘protein leverage’ (PL) [9–10]: food is consumed until absolute intake of protein meets (or approaches) a target value, regardless of the consequences (i.e., shortfall or overconsumption) for other nutrients in the diet and TEI. Evidence for PL in humans has been observed in several different contexts, including tightly controlled experiments in lean humans [11], in humans on high-protein diets [12], in a free-living human population [13], in a cohort of youth with obesity [14], and in quantitative syntheses of published data [15, 7].

An inevitable consequence of PL is that, as the amount of protein within a food/diet falls, net food intake must increase to meet the protein target. Where protein is diluted by an energy yielding nutrient (e.g., carbohydrate or fat), PL will lead to excessive energy intake. The PL hypothesis (PLH) posits that PL combined with a dilution of dietary protein by carbohydrates and energy-rich fats in modern (largely western) food supplies is a driving factor behind the world’s obesity epidemic [4, 7, 14].

Most studies on PL to date have concerned younger and active people. However, the world counted 703 million older persons in 2019, and it is projected in 2050 that one in six people in the world will be aged 65 years or over [16]. It is not evident that the relationships between protein intake (through PL), TEI, and body mass index (BMI) observed in younger adults would hold in older adults. Indeed, total daily energy expenditure changes substantially with age and declines especially for older adults [17]. Furthermore, in mice, high protein intake appears to be associated with higher mortality in younger mice, but is protective in older mice [18]. Likewise, in humans, the relationship between TEI and BMI is weak in some cases [19] and nothing is known about this relationship in older adults. In this study, we considered a cohort of individuals aged 67-84 years who were enrolled in the Quebec Longitudinal Study on Nutrition and Successful Aging (NuAge) [20]. The aim of the study was to test for PL and signatures consistent with the PLH in this cohort of older adults.

## METHODS

### Study cohort

We used data from the NuAge study [20]. NuAge is a longitudinal study that aimed to assess the effect of diet and nutrition on health in older adults. Participants were selected randomly from the Quebec Medicare database after stratification for location (Montreal, Laval, Sherbrooke areas in Quebec, Canada), age (three age groups: 67-72, 73-77, 78-84) and sex (men and women). The sample consisted of 1793 community-dwelling persons (1587 recruited and 206 volunteers) in good health at recruitment in 2003-2004 (T1). Participants were re-examined annually for three years (T2, T3 and T4). Subsequently, 1753 of these participants consented to the incorporation of their data into the NuAge Database and Biobank, used here. The study and the NuAge Database and Biobank were approved by the Ethics Committee of the CIUSSS-de-l’Estrie-CHUS (Quebec, Canada). Here, we considered the cohort T1 (retrospective analysis), except as otherwise indicated.

### Dietary intake data

Intakes of macronutrients were assessed using three non-consecutive 24-hour dietary recalls (of which one was during a weekend) using the USDA 5-step multiple-pass method [21]: 1 face-to-face and 2 telephone interviews [20]. These interviews were conducted by trained research dietitians, who used graduated utensils and photos of standardized food portions to enhance portion-size estimation [22]. Only macronutrients coming from foods (i.e., carbohydrates, protein and fat) and participants with at least two 24-hour dietary recalls were considered. Macronutrient intakes were computed using the CANDAT-Nutrient Calculation System (version 10, ©Godin London Inc.) based on the 2007b version of the Canadian Nutrient File from Health Canada and a database of > 1200 additional foods [23]. The mean daily macronutrient intakes from two to three 24-hour recalls were the data used in these analyses. Participants with at least one missing observation of macronutrient mean intake, height, or body weight were excluded. The sample size was therefore reduced to 1,699 participants. Other variables considered for sensitivity analyses (using stratified analyses) are the sex, age group (as stated above with the recruitment procedure), self-reported diabetes status [24], smoking status [25], and physical activity level using the Physical Activity Scale for the Elderly (PASE) [26].

### Statistical data analysis approach

All analyses were performed using version 4.1.1 of R software [27] and were independently replicated by a second analyst to avoid bias and to ensure accurate results.

### Determination of strength of leverage

Based on the mean daily protein, carbohydrate, and fat intake expressed in grams/day (*gP*, *gC* and *gF*) for each participant, we computed the energy intake from each macronutrient. We estimated TEI using equation (Eq. 1) below and then we calculated the energy contribution (EC; i.e. % of energy) of each macronutrient to the TEI. BMI, i.e., the ratio between measured weight (kg) and squared measured height (m), was calculated. In addition, scatter plots were used to visualise the relationship between TEI and macronutrient contributions alone and in pairs by age group and sex.

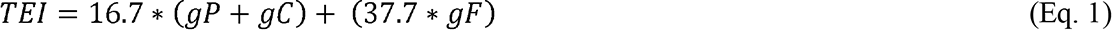

The Gauss-Newton algorithm was used to determine the weighted least square estimates of the parameters of the power function (Eq. 2), to predict the strength of leverage from the relevant EC from macronutrients toward the TEI [14].

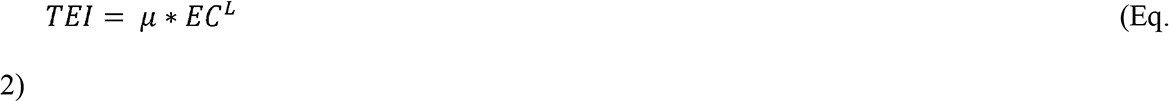

where µ represents the mean energy intake of a diet composed of 100% of the macronutrient (expressed in kJ) observed in the cohort, and *L* is the strength of leverage. Here, complete leverage, where the intake is regulated to reach µ regardless of the consequences for total energy intake, would manifest as *L* = −1. In contrast *L* = 0 indicates null macronutrient leverage, in that the modelled nutritional component and its reciprocal are equally strongly regulated [7, 14]. Positive values (i.e. *L* > 0) indicate that increasing energy from a given macronutrient is associated with increased TEI; because percentage intake of the three macronutrients sums to 100%, this would generally be detected as the converse of leverage (L < 0) in another macronutrient, and would not necessarily be of particular interest in and of itself.

In addition to running analyses on raw TEI, we generated a height-weight adjusted TEI by linearly adjusting TEI in an additive-effects model of weight and height from which we extracted the residuals (“adjusted-TEI”). Linearity of the relationships and normality of the residuals were both verified. Power models were also run on adjusted-TEI to evaluate the combined effect of weight and height on the PL strength when compared to unadjusted TEI. The adjusted TEI adjusts for the joint effect of height and weight (without the variance in TEI associated with weight and height). Therefore, if PL becomes stronger (i.e., *L* is more negative) with adjusted TEI in comparison to unadjusted TEI, it would suggest that the PL effect is particularly robust.

Sensitivity analyses were run on population subgroups to assess the generality of findings (Table S1). We considered two age groups (the youngest: 67-72 and the oldest: 78-84, in the cohort), sex (men and women), three BMI groups [low: BMI <=22, normal: 22<BMI<=27, high: BMI > 27], two groups based on reported diabetes status (non-diabetic and diabetic: all types), and three smoking groups (never-smoker, former smoker, current smoker). PASE scores were used to categorize participants’ physical activity levels into three groups based on quartiles 1 and 3 (PASE < Q1; Q1 <= PASE <= Q3, PASE > Q3). Finally, we also evaluated the link between TEI at a time *t* (e.g., T1) and the gain in body mass reflected by the difference in BMI between time points *t* and *t*+1 (e.g., T2-T1).

### Compositional modelling of macronutrients contributions toward TEI

Modelling the TEI considering the additive effect of the EC from all three macronutrients requires the consideration of the linear correlation and mutual constraints existing between these contributions (∑_J_ EC_J_ = 1, the main feature of mixture experiments). Failure to consider this correlation when fitting models to mixture experiments can lead to inappropriate estimation of the residual variance and potentially inappropriate inference from the inevitably correlated model coefficients [28]. We fitted five models: a null linear model and the first four Scheffé models [29] (see supporting information for more details) using the R package *mixexp* [28]. We selected best-fitted models using AIC [30], where the model with lowest score is favoured. Predictions from the AIC-favoured model were then plotted by projection as a contour plot on a right-angle mixture triangle [31]. The same models were performed on adjusted-TEI and BMI.

Additionally, we noticed that the contributions of fats and carbohydrates were strongly negatively correlated (Figure S1). This strong negative association between %C and %F means that the informative information is bivariate along two largely independent axes: %P on the one hand, and %carbohydrates:%fat (CF) balance on the other. Thus, as a more intuitive but less formally correct alternative to mixture models, we used principal component analysis (PCA) on EC from fat and carbohydrate to create a new variable (transformed CF), which explains 93% of the total variance in fat and carbohydrate intake (Figure S2) and is qualitatively similar to a CF ratio. The function *princomp* from the built-in R package *stats* [27] was used for this purpose. The new variable was used as a predictor with the EC from protein in linear models to assess their effects on TEI, adjusted-TEI, and BMI.

## RESULTS

### Cohort features: macronutrient intake, TEI and BMI

Carbohydrates contributed to the TEI of older adults more than proteins and fats (Table 1 and Figure S3). Generally, men aged 73–84 years (age groups 2 and 3) had the highest TEI, and the mean TEI of men (8 466 ± 2 113 kJ) was greater than women (6 946 ± 1 719 kJ). However, 4% of the youngest (67–72 years) recorded a TEI > 12 000 kJ compared to 2% of the oldest (78–84 years). Most participants with higher EC from protein (EC > 25%) recorded lower TEI (Figure 1). The median TEIs for women, men, and the whole cohort were 6 780, 8 270, and 7 442 kJ, respectively. EC from carbohydrates and fats present inverse trends on TEI (Figure S1). Higher EC from carbohydrates is related to lower EC from fat and *vice versa*, but the highest values of TEI were observed for EC from fats ranging between 25 and 45 % and for EC from carbohydrates ranging between 40 and 60 % (Figure 1). The mean BMI of all participants is 27.89 and it can be noted that regardless of age group and sex, the mean BMI is above 27 (the threshold for obesity in older adults). In fact, 908 of the 1 699 participants considered (53.44%) had obesity, and 1.65% participants had severe cases (BMI > 40) (Table 1).

**Figure 1.**
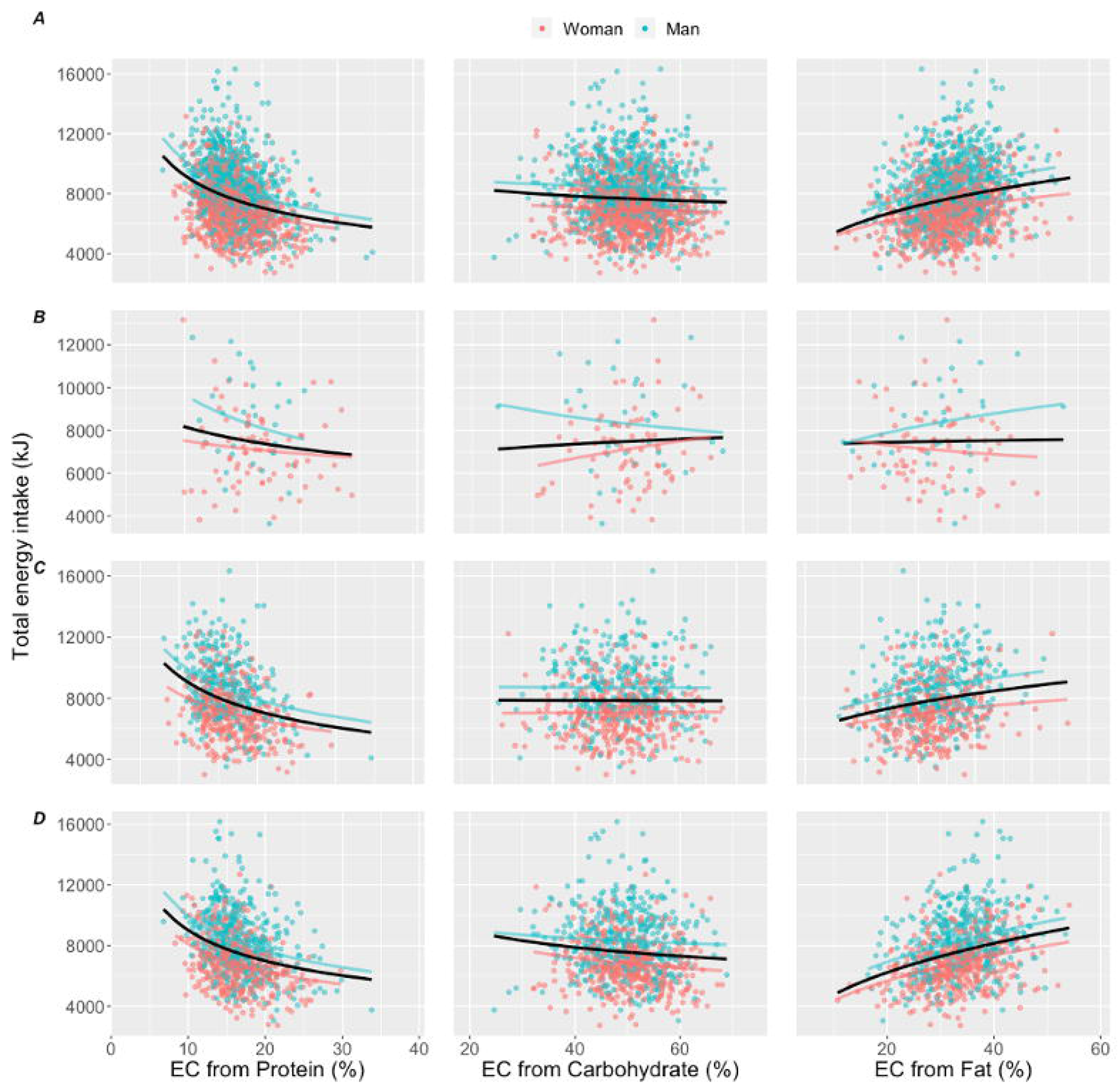
Trends of TEI according to energy contribution (EC) for each macronutrient (%) with differentiation of individuals by their sex for (**A**) all individuals, (**B**) individuals with low BMI (BMI ≤ 22), (**C**) individuals with normal BMI (22 < BMI ≤ 27) and (**D**) individuals with high BMI (BMI > 27). Curves represent power law functions for each sex (red and blue for women and men, respectively) and joint (black).

### PL in older adults

There was evidence for protein leverage on TEI, indicated by a negative *L* coefficient in power regressions (-0.37 in the whole cohort), which was statistically significant globally and in all sub- populations (*p* ≤ 0.001 in most models; Figures 1 and 2). There was also a negative *L* coefficient for dietary carbohydrates, although this was comparatively weak, ranging from -0.19 to -0.08 depending on the subset. The *L* coefficient for carbohydrates was statistically significant for High BMI, individuals without diabetes, and when considering the whole cohort, although only marginally so (p = 0.048). This suggests a weaker leverage for carbohydrates than for protein. Fat intake, by contrast, was positively associated with TEI (0.31 in the whole cohort) and highly significant in most models (Figures 1 and 2). This is to be expected in a mixture setting where leverage for protein and a weaker effect of carbohydrates together drive passive intake of fat. PL was stronger in some subgroups than others (Figure 2): the current smoker group (-0.66), the older age group (-0.47), the group with diabetes (-0.43), the lowest physical activity quartile group (-0.42), the normal BMI group (-0.40), and the group with obesity (-0.37). PL strengths were the same for participants (i) with diabetes and lower PASE, and (ii) without diabetes and with higher PASE (Figure 2). An interesting result here is the marked increase (absolute value) of PL after adjusting TEI for weight and height. For example, in the full cohort, the strength of protein leverage increases from -0.37 to -0.54 when adjusting for height and weight, and in current smokers, it increases from -0.66 to -1.09. This effect was observed for most subgroups (Figure 2, Table S1).

**Figure 2.**
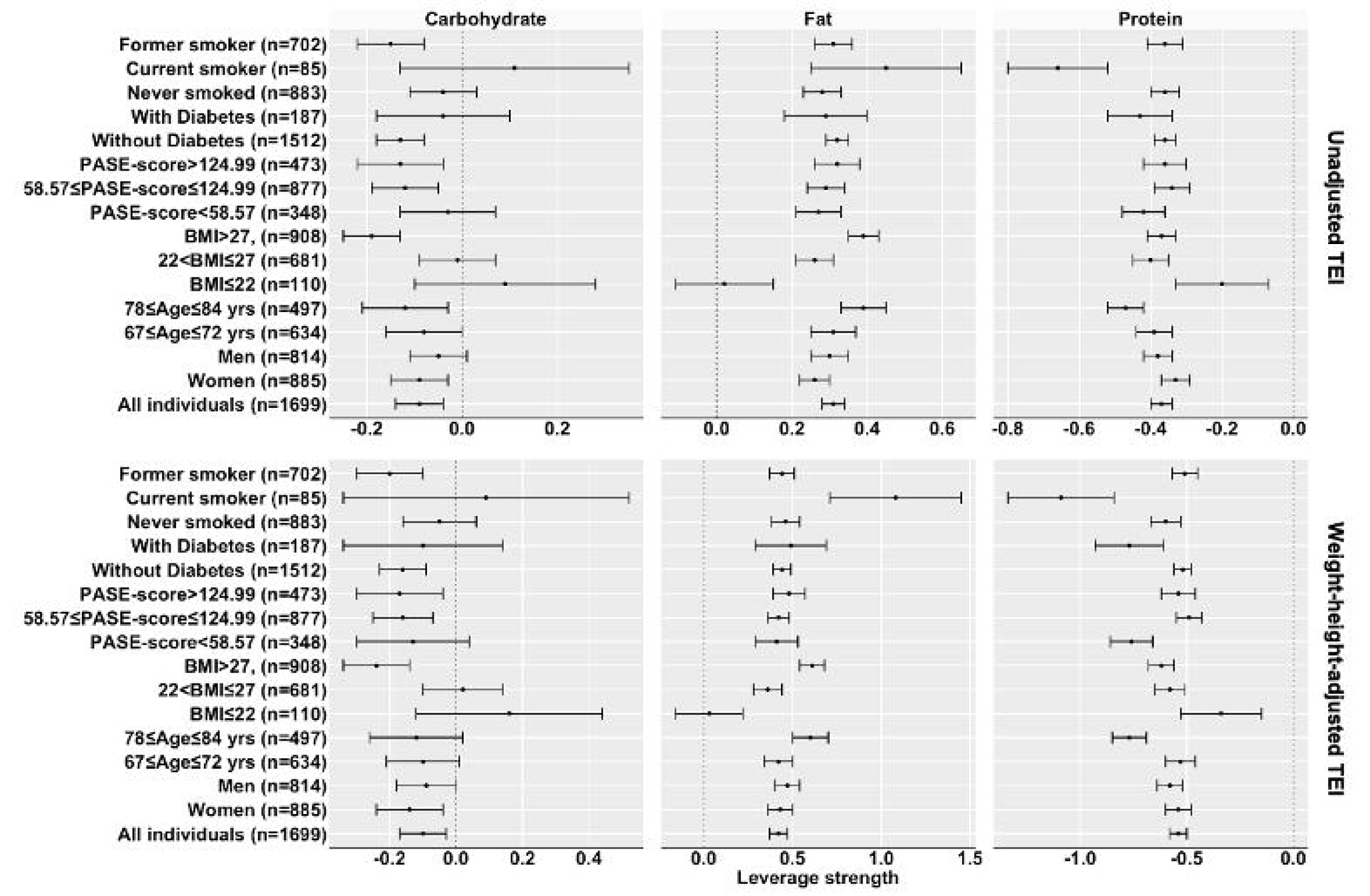
Nonlinear power regression estimates of macronutrients strength of leverage regarding unadjusted total energy intake (TEI), weight-height-adjusted TEI and body mass index. Each graph has three panels of forest plots (for the three macronutrients), and each line represents the estimated strength of leverage with its confidence interval.

In contrast to the leverage findings, BMI and TEI were completely unassociated with each other, globally or when stratified by years of follow-up or age group (Table S4, Figures S5 and S6). This appears to indicate a nearly complete uncoupling of TEI and BMI within this cohort of older adults. Besides, the lack of relationship when stratified by BMI shows that the relationship is not simply non-linear. Overall, increases and decreases in both BMI and TEI over time (visits) were relatively balanced: Figure S6B shows scatterplots symmetrical around no change in either variable.

### Optimal macronutrient composition to meet TEI requirements

Model 2 (first Scheffé’s mixture experiment model; see supplementary material for more details), which assesses the linear effects of all ECs from each macronutrient, had the most favourable AIC among all models when testing for effects of EC on TEI, adjusted-TEI, or BMI (Figure S7, Tables S2 and S5). For all individuals and age group 67-72 years old, a combination of moderate EC from carbohydrates and/or fats (40 - 60 %) and a low EC from proteins (10 – 20 %) provided higher TEI, as shown by the model predictions represented by right-angle mixture triangles (Figure 3). These projections clearly show that the principal gradients in TEI and weight-height adjusted-TEI are along the protein axis, not the fat or carbohydrate axes (Figure 3A-B) while the principal gradient in BMI is along the carbohydrate and fat axes (Figure 3C). Considering all individuals, each percent of energy intake from proteins reduced TEI by 77 kJ on average, *ceteris paribus* (Table S2). Increasing carbohydrates and/or fats at the expense of protein increases TEI passively. This scenario was similar for age groups 67-72 and 78-84 years (Figure S7). This result was also replicated using the PCA-derived EC from carbohydrate-to-fat variable (“transformed CF”), which is qualitatively similar to a carbohydrate:fat ratio. For all age groups, TEI strongly decreased as a function of the transformed CF, implying that for a given level of EC from protein, TEI is higher in individuals who get a larger percent of their remaining calories from fat rather than carbohydrates (Figure S8), likely due to the weak leverage of carbohydrates (Figure 2).

**Figure 3.**
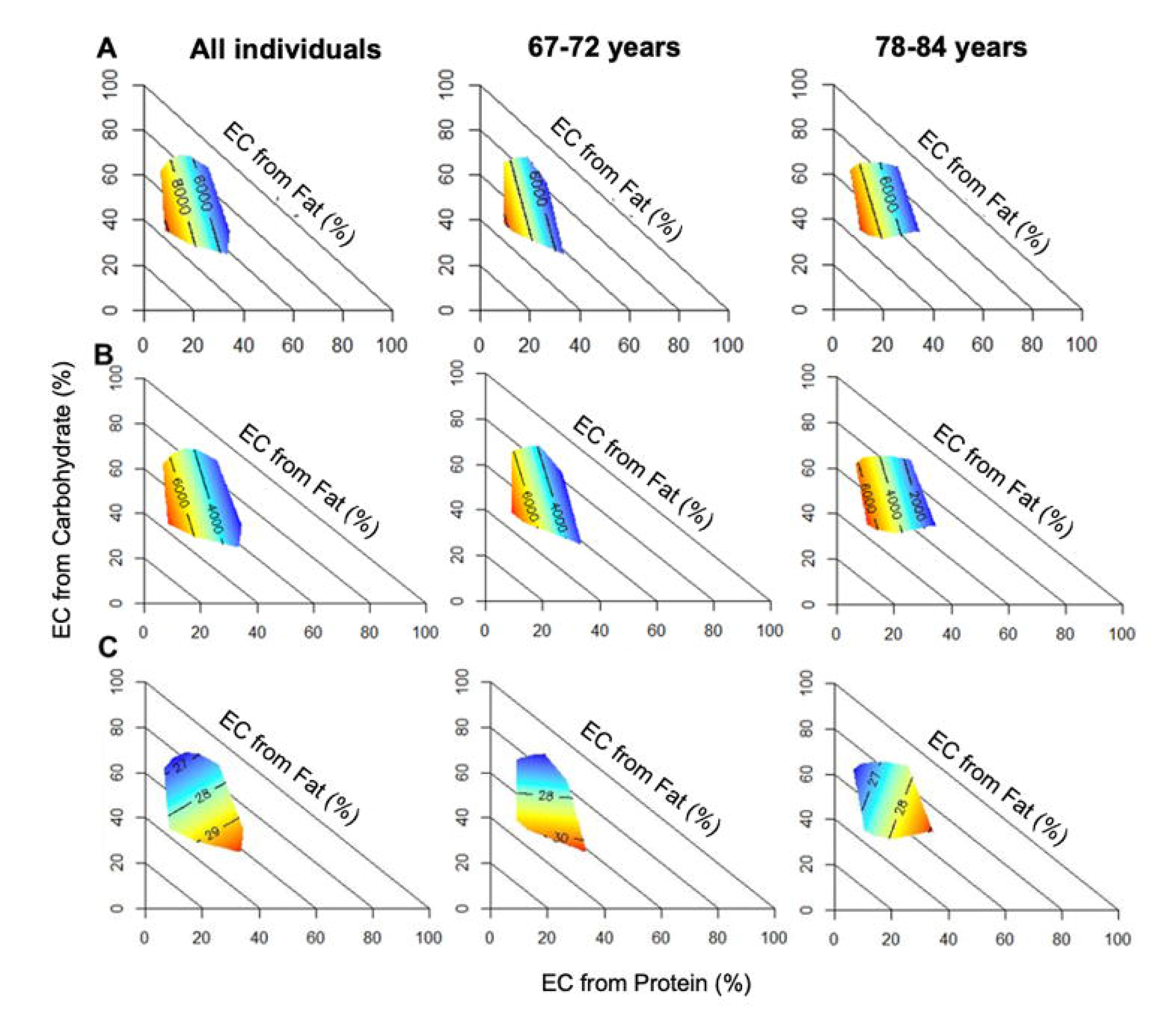
Right-angle mixture triangles showing effects of energy contribution (EC) from each macronutrient on (**A**) total energy intake (TEI), (**B**) height and weight adjusted-TEI and (**C**) body mass index (BMI) in all individuals and age-group subsets (67-72, 78-84 years). Surfaces indicate values of TEI (kJ), adjusted-TEI (kJ) or BMI (kg/m^2^), where blue is the minimal value on the surface and red is the maximal value.

## DISCUSSION

In this retrospective study, we tested for and found extensive evidence of PL in older adults (*L* = -0.37 globally). This leverage is stronger when TEI is adjusted for the weight/height of individuals (*L* = -0.54) and is broadly replicated in subpopulations and various sensitivity analyses. This result is similar to findings by Saner et al. [14], who also showed evidence for PL in overweight youth. However, we detected no significant effect of percentage energy from protein on BMI as predicted by the PLH, likely because TEI and BMI appear to be uncoupled in this population.

### No evidence that protein leverage affects BMI in this healthy aging cohort

In this cohort of healthy older adults, we did not detect signals consistent with PLH (e.g., an effect of energy from protein on BMI). This result however is not necessarily surprising because we found that TEI and BMI are uncorrelated (no clear trend for TEI or BMI over time, nor any association between the two); PLH posits that the leverage of protein on TEI will translate to an effect on BMI because excess energy intake increases adiposity. This uncoupling could be because of the short-term timescale over which dietary intakes were measured compared to the long duration over which changes in body mass accumulate. In another perspective, it can reflect a gradual adaptation of the system to lower caloric intake by reducing energy expenditure [32]; the body requires less energy to ensure the system’s basic functionalities [33]. Accordingly, metabolism can become more efficient, allowing the body to survive with less energy than bodies of similar weight that have not been deprived of calories [34].

However, we could also question whether BMI is indeed a fair metric of PLH in older adults. For instance, sarcopenia, a chronic disease prevalent in older adults, is a progressive loss of muscle mass and strength which is related to age, sex and levels of physical activity [35–36]. Therefore, a low or decreasing BMI may be associated with the presence of sarcopenia [37–38]. Because both sarcopenia and obesity may coexist within an individual and confound each other via BMI, BMI is increasingly imprecise with advancing age, and DEXA fat mass might be a more relevant metric for PLH in future studies.

Along these lines, we thus hypothesized that high protein intake could prevent weight loss in the context of low BMI, but also prevent weight gain in the context of high BMI (obesity). We therefore stratified the analyses by BMI, predicting a positive relationship between protein EC and BMI in the low BMI subgroup, and a negative relationship between protein EC and BMI in the high BMI subgroup (Figure S4). There was no evidence for such effects, and if anything, we observed the inverse: a hint of protein leverage on BMI in the lowest BMI stratum.

### Protein leverage in smokers

In this cohort of older adults, current smokers recorded the strongest PL, more than 50% higher than in the full cohort (L=-1.09 vs. -0.66 in the weight-height adjusted analyses). Despite the small proportion of this subgroup in the study sample (85 individuals; 5%), this finding is noteworthy as it points to a potential association between PL and smoking. Indeed, this increase in PL might be associated with high concentrations of the fibroblast growth factor 21 (FGF21). FGF21 is a liver-derived peptide hormone that stimulates glucose uptake and lipid metabolism [39] and regulates protein phosphorylation and insulin sensitivity [40]. Recent studies have shown a significant and positive association between regular smoking and high concentrations of FGF21 [41–42]. In addition, an evaluation of FGF21 levels in humans showed differential expression for people who smoked every day. They also showed its role in the regulation of sweet food intake in adults, which could affect nutrient-specific appetite [42]. Furthermore, FGF21 is upregulated in protein deprivation and obesity, supporting a possible association between PL and FGF21 [43].

### Limitations and strengths of the study

Nonlinear modeling is challenging because it requires identifying the best nonlinear function to fit the data. In this study, we opted for the power law function, which is not necessarily proven to be the best nonlinear function for our data. We have not investigated other nonlinear functions. However, the power function is the most used in recent studies on PL and PLH [14]. Moreover, other alternatives to Scheffé’s mixture model (used in this study) can be investigated. The nonlinear structural equation mixture approach, which integrates the specification of the nonlinear function and the flexibility of semiparametric structural equation mixture approaches for approximating the nonnormality of latent predictor variables, may be worth investigating [44]. Further, the isometric log-ratio function, which is an intuitive approach to stabilize imbalance weight distributions in mixture experiments, is also a possible approach to explore for mixture experiments analyses [45].

This study focused on a population of community dwelling older adults who reside in Quebec and were generally healthy at recruitment [20]. This population is relatively homogeneous. Given the diversity of lifestyles of different populations around the world, we are aware that this cohort is not representative of older adults in general. The study also considered four follow-up time points over ∼3 years, a follow-up that may be insufficient. More data over a longer period could allow us to observe relevant patterns between TEI and BMI. However, associations between TEI and BMI are weaker over T1-T4 than over a one-year interval (e.g., T1-T2; Table S4), so it is unlikely results would be stronger with longer follow-up.

While some observational designs, combined with appropriate statistical methods, can give strong inferences on causality, this study was not designed for such inferences, and cannot make claims about causality. Nonetheless, the clear confirmation of theoretical predictions for specific functional relationships, consistent with findings in other cohorts and in animal models, lends credence to our findings as representing broad patterns of interest.

Despite these limitations, this study has several strengths and is generalizable to older adults in other Canadian provinces and Western countries. It used data from the NuAge study [20], which has a large sample. We used state-of-the-art statistical methods to analyze macronutrient compositions. Intakes of macronutrients were assessed using three non-consecutive 24-hour dietary recalls (of which one during weekend) using the USDA 5-step multiple-pass method [21]: one face-to-face and two telephone interviews [20]. This approach is also a state-of-the-art quantification method of dietary intake in an observational context. Additionally, the interviews were conducted by trained research dietitians [22].

## CONCLUSIONS

This study assessed evidence for PL in adults aged between 67 and 84 years. We showed evidence for PL on TEI in the cohort, but no association between TEI and BMI was detected, making tests of PLH irrelevant. TEI and BMI thus appear to be decoupled in this cohort, in contrast to most younger cohorts. The confusion may reflect the weaker and more complex relationship of BMI and health status in older adults. These results are consistent with the hypothesis that eating more protein during aging can prevent muscle loss and provide the required energy to prevent weight loss [22, 43] and sarcopenia, which is a disorder that is becoming increasingly prevalent among people with obesity in older age [35]. PL was stronger for older adult men, people with diabetes, current smokers, and those with higher BMI or lower physical activity, suggesting that these groups would be even more favored to increase their protein intake to control their body weight and preserve their muscle mass.

## AUTHOR CONTRIBUTIONS

SHH conducted data analysis data and wrote the manuscript. AAC designed the study and reviewed the manuscript. NP, VT gave access to NuAge data and reviewed the manuscript. VL, PG, SJS and DR reviewed the manuscript.

## DATA AVAILABILITY

All data supporting the conclusions of these analyses are presented in the manuscript or the supplementary material. Details of additional data can be obtained from the study authors upon reasonable request.

## Data Availability

All data produced in the present study are available upon reasonable request to the authors.

