## Supplemental information for "Evidence for protein leverage on Total Energy Intake, but not Body Mass Index, in a large cohort of older adults"

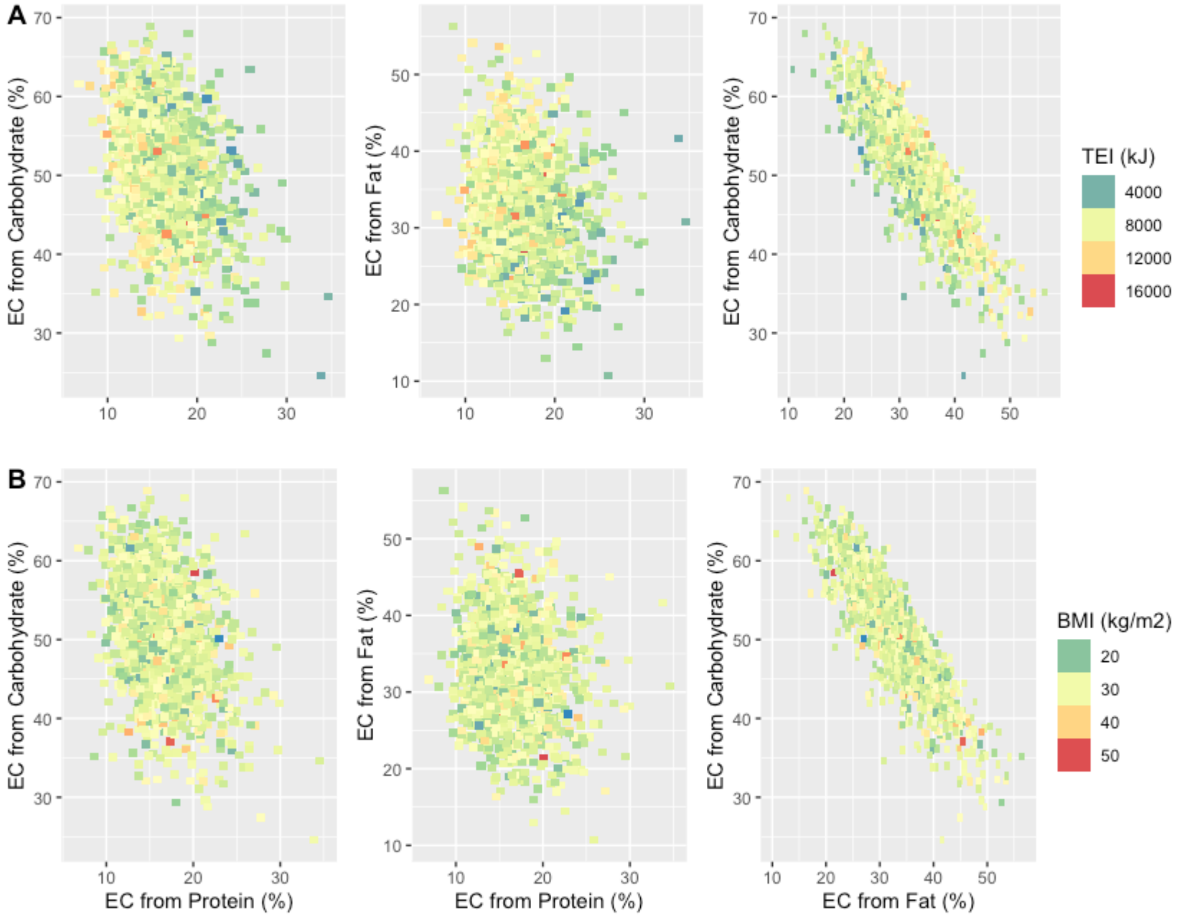


**Figure S1**. Distribution pattern of (**A**) unadjusted TEI and (**B**) BMI of participants in relation to the EC (%) from protein and carbohydrate (left), protein and fat (middle), and fat and carbohydrate (right), for all individuals.

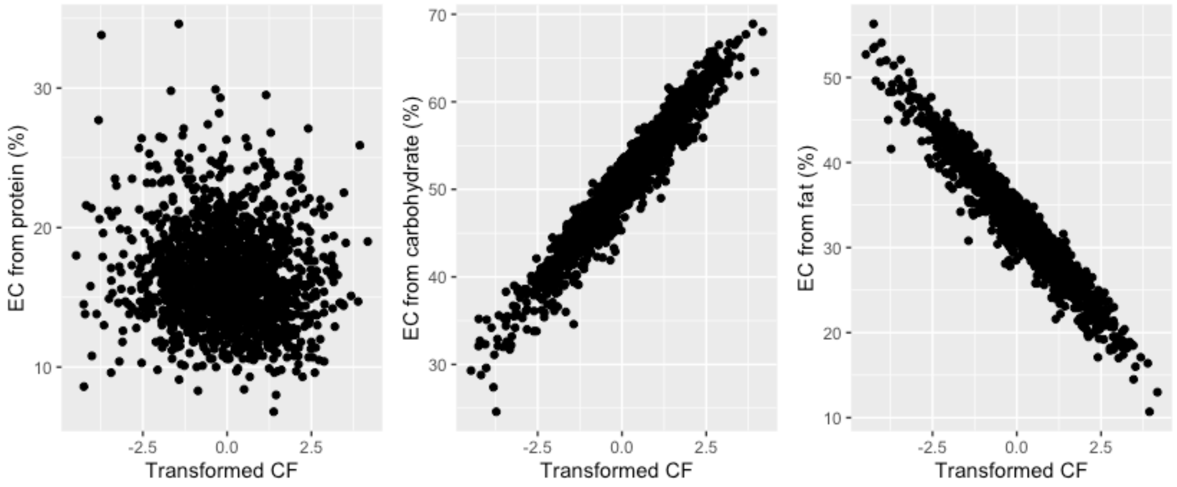


**Figure S2**. Trends between the new PCA-derived variable (transformed CF; from carbohydrates and fat EC) and EC from each macronutrient (%).

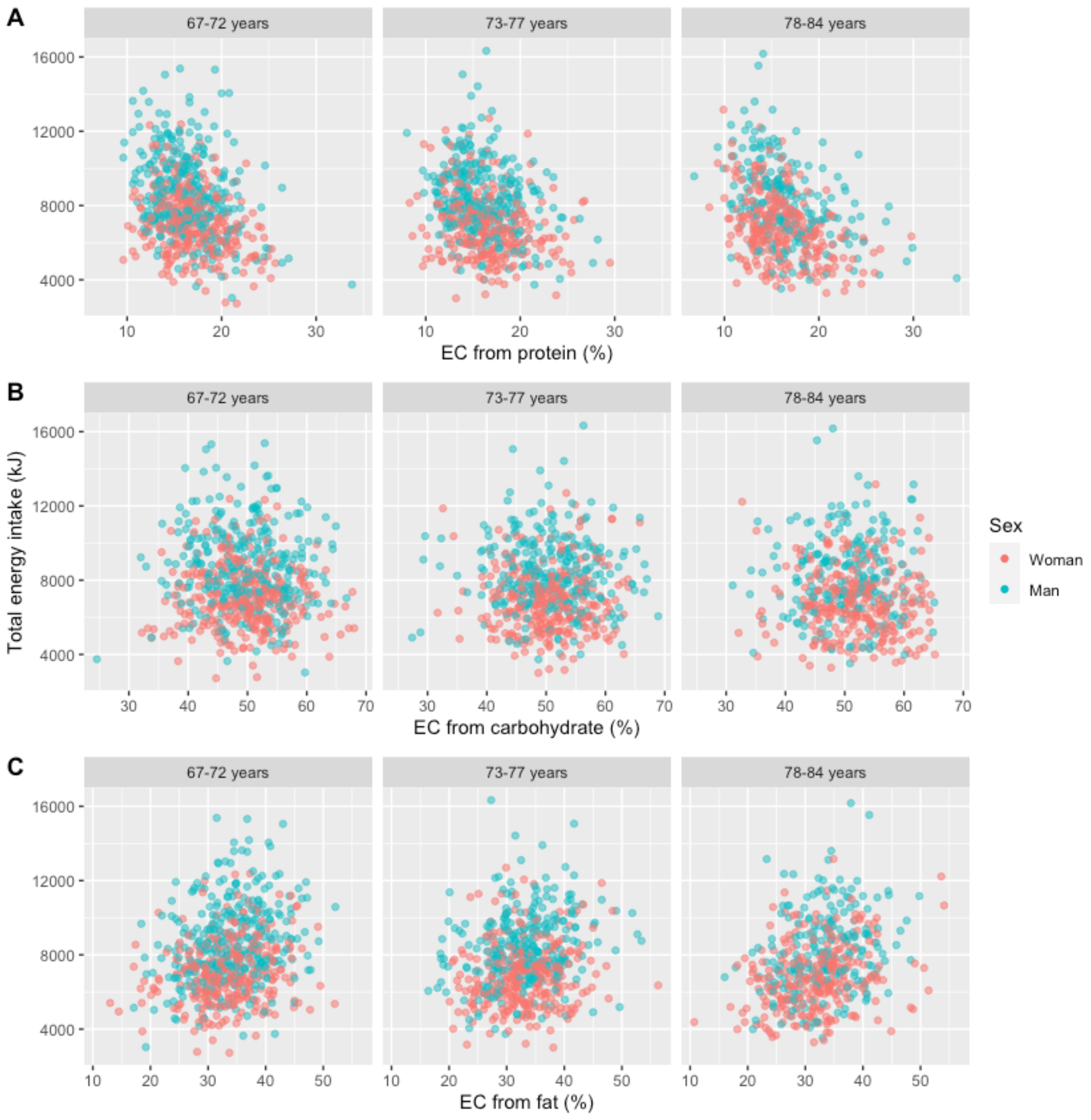


**Figure S3**. Overview of relationships between unadjusted TEI and (**A**) energy contribution (EC) from protein (%), (**B**) energy contribution (EC) from carbohydrate (%) and (**C**) energy contribution (EC) from fat (%) according to age groups of individuals and their sex.

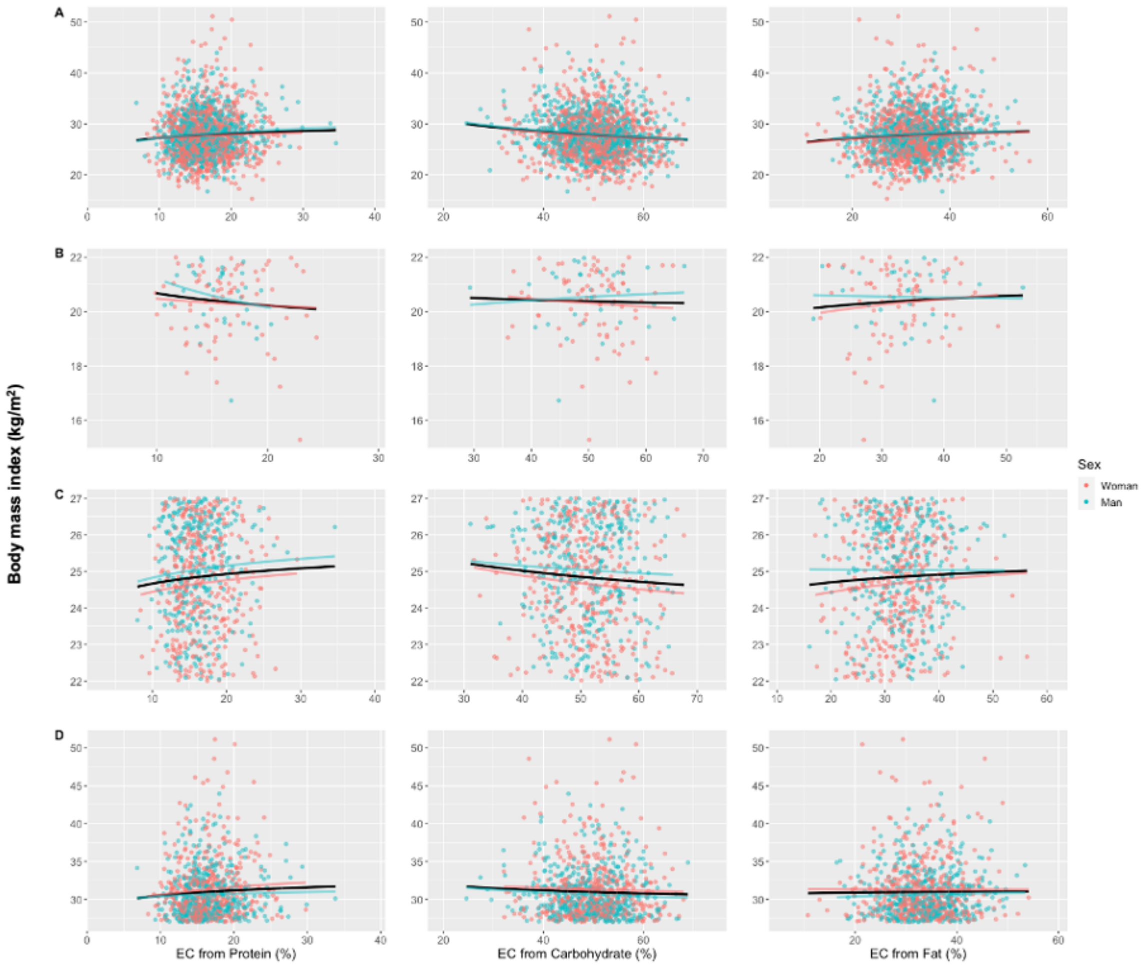


**Figure S4**. Trends of BMI according to energy contribution (EC) for each macronutrient (%) with differentiation of individuals by their sex for (**A**) all individuals, (**B**) individuals with low BMI (BMI ≤ 22), (**C**) individuals with normal BMI (22 < BMI ≤ 27) and (**D**) individuals with high BMI (BMI > 27). Curves represent power law functions for each sex (red and blue for women and men, respectively) and joint (black).

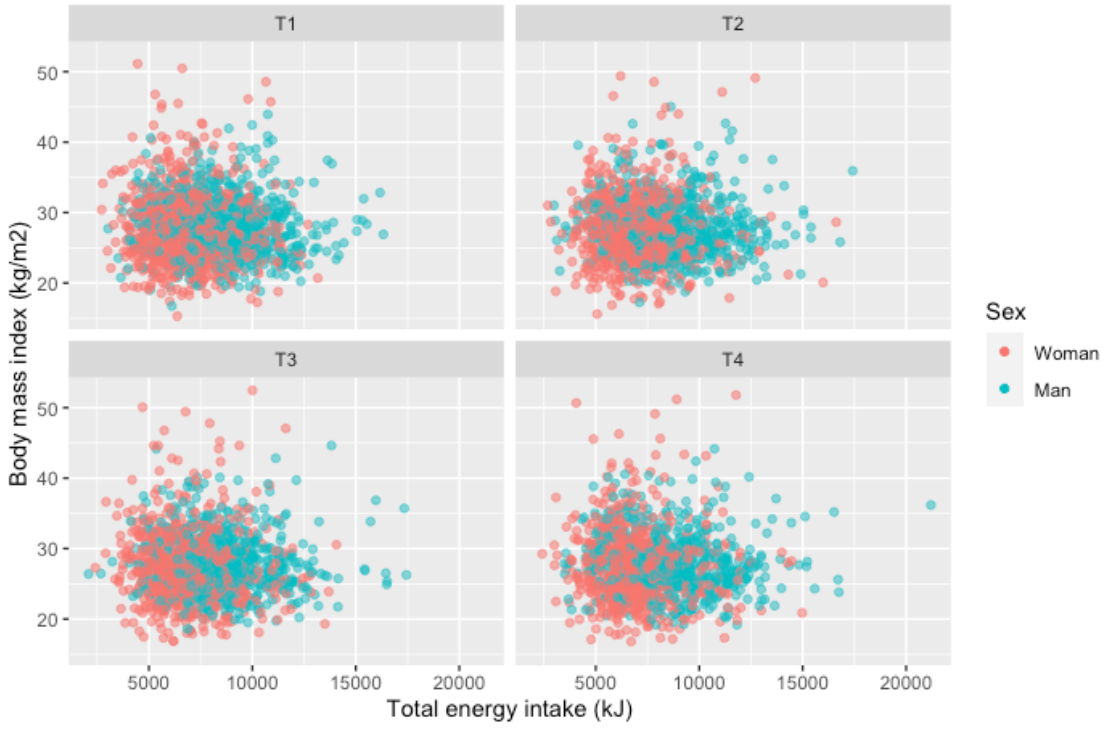


**Figure S5.** Scatter plots showing associations of total energy intake with body mass index at each annual visit (T1 to T4) for all individuals.

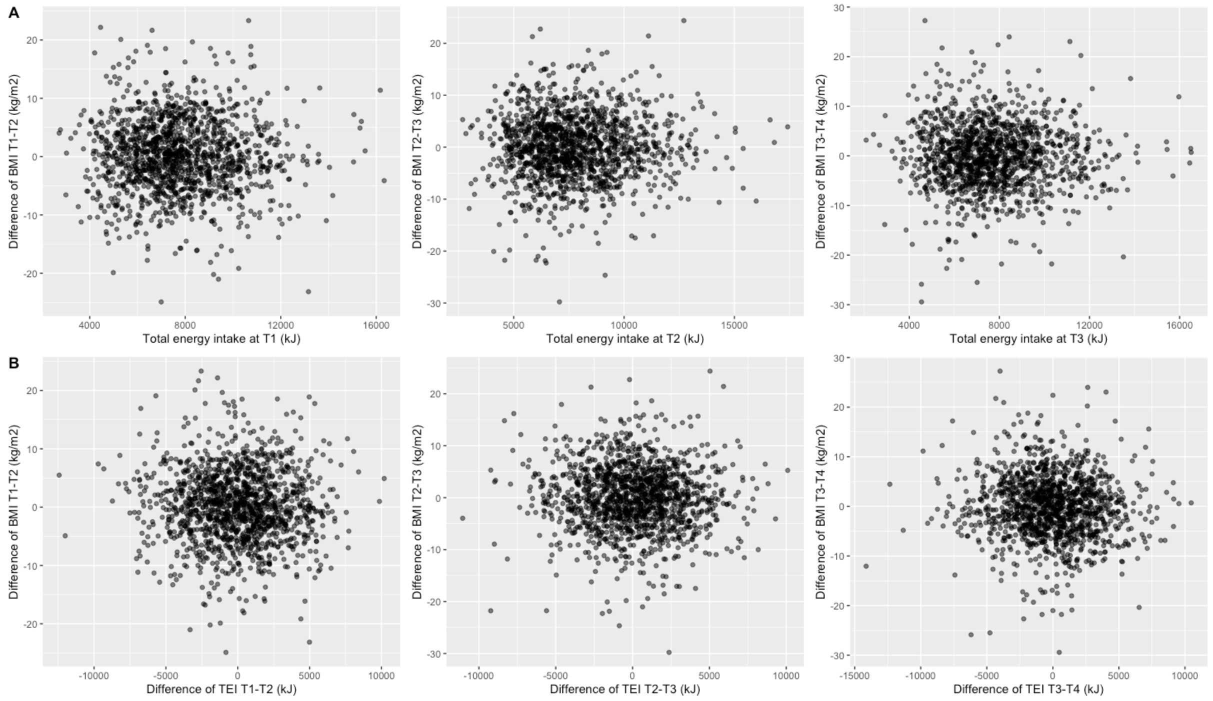


**Figure S6.** Scatter plots showing (A) associations of total energy intake at T1, T2 and T3 with the difference of body mass index between T1 and T2, T2 and T3, T3 and T4, respectively; (B) associations of differences in TEI and BMI between visits T1 and T2, T2 and T3, T3 and T4.

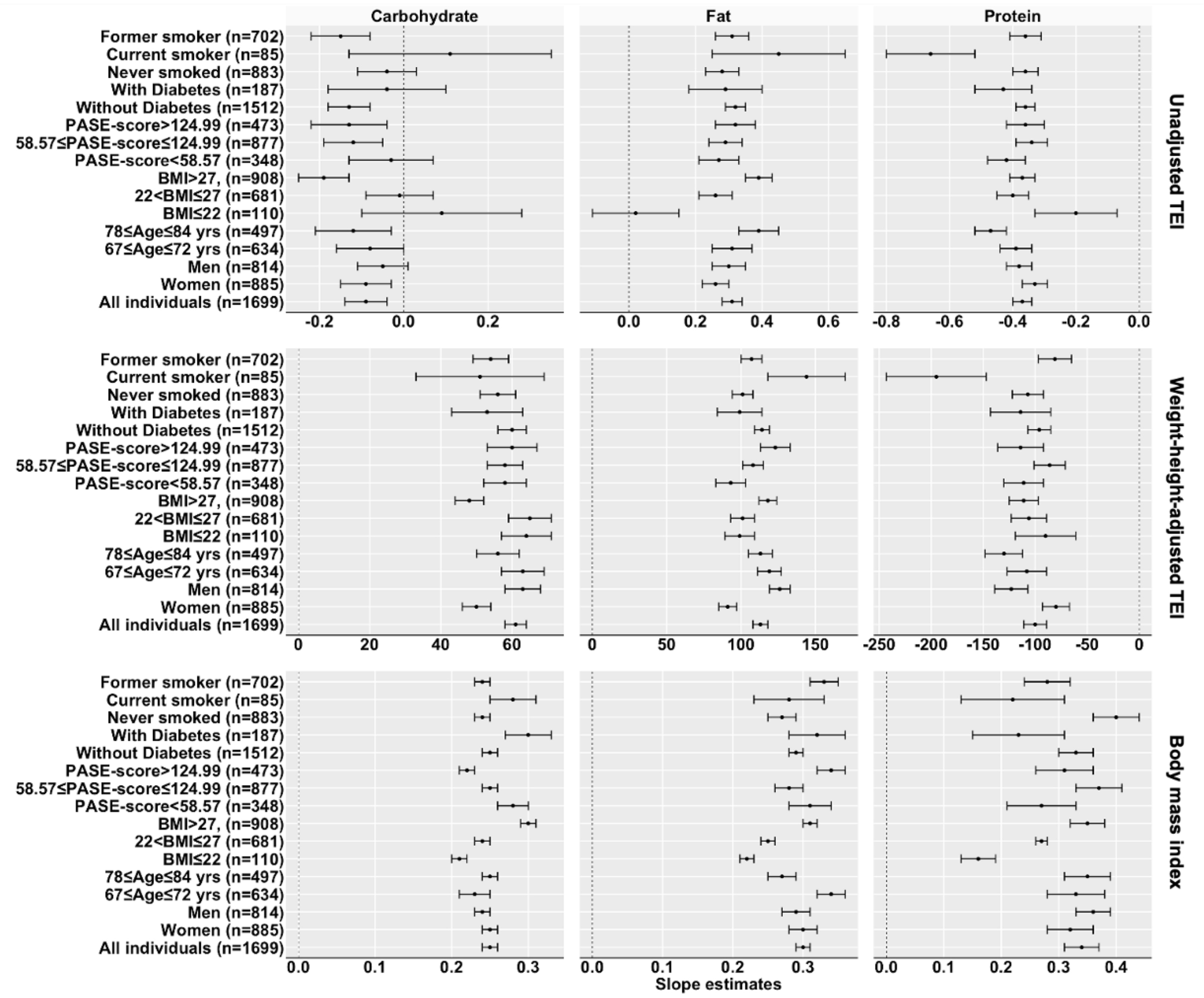


**Figure S7.** Mixture experiment model estimates of macronutrient effects on unadjusted total energy intake (TEI), weight-height-adjusted TEI and body mass index. Each graph has three panels of forest plots (for the three macronutrients), and each line represents the estimated slope with its confidence interval.

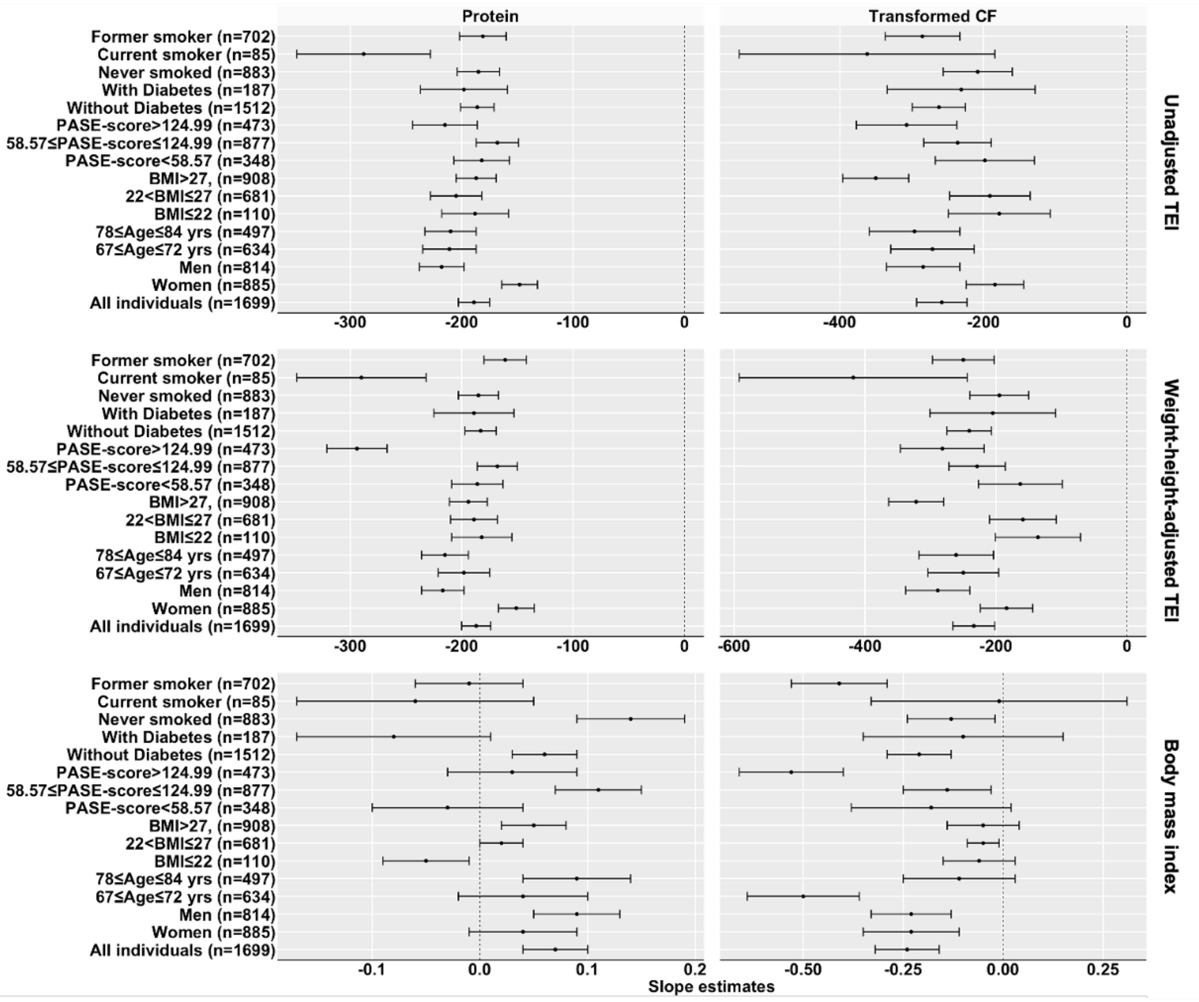


**Figure S8**. Ordinary least squares estimates of protein and the PCA-derived “transformed CF” effects on unadjusted total energy intake (TEI), weight-height-adjusted TEI and body mass index. Each graph has two panels of forest plots (for protein and for PCA-derived “transformed CF”), and each line represents the estimated slope with its confidence interval. Of note, the PCA-derived “transformed CF” is qualitatively similar to a carbohydrate:fat ratio.

**Table S1**. Results from power functions fitted between macronutrients and TEI and BMI: target intake μ (mean energy intake of a diet composed of 100% of the macronutrient) and strength of leverage (*L*) with their standard errors (se) and p-values (** for $p \le 0.001$, * for $0.001 < p \le 0.05$ and none for $p > 0.05$)

| EC from macronutrients | TEI | | Weight-Height adjusted-TEI | | Weight adjusted TEI | | BMI | |
|---|---|---|---|---|---|---|---|---|
| | $\mu.10^4 \pm se.10^4$ | *L* ± se | $\mu.10^4 \pm se.10^4$ | *L* ± se | $\mu.10^4 \pm se.10^4$ | *L* ± se | μ ± se | *L* ± se |
| **All individuals (n = 1,699)** | | | | | | | | |
| Protein | 2.14±0.18** | -0.37±0.03** | 2.28±0.26** | -0.54±0.04** | 2.41±0.29** | -0.56±0.04** | 24.62±1.31** | 0.04±0.02* |
| Carbohydrate | 1.12±0.21** | -0.09±0.05* | 0.77±0.20** | -0.10±0.07 | 0.73±0.20** | -0.09±0.07 | 42.13±4.70** | -0.11±0.03** |
| Fat | 0.26±0.03** | 0.31±0.03** | 0.12±0.02** | 0.42±0.05** | 0.12±0.02** | 0.43±0.05** | 23.70±1.64** | 0.05±0.02* |
| **Women (n = 885)** | | | | | | | | |
| Protein | 1.72±0.19** | -0.33±0.04** | 1.87±0.32** | -0.54±0.06** | 1.80±0.31** | -0.52±0.06** | 25.42±2.08** | 0.03±0.03 |
| Carbohydrate | 0.98±0.24** | -0.09±0.06 | 0.72±0.29* | -0.14±0.10 | 0.73±0.30* | -0.14±0.10 | 40.65±7.20** | -0.10±0.05* |
| Fat | 0.28±0.04** | 0.26±0.04** | 0.09±0.02** | 0.43±0.07** | 0.10±0.02** | 0.43±0.07** | 23.58±2.47** | 0.05±0.03 |
| **Men (n = 814)** | | | | | | | | |
| Protein | 2.45±0.27** | -0.38±0.04** | 2.69±0.45** | -0.58±0.06** | 2.82±0.49** | -0.60±0.06** | 23.68±1.59** | 0.06±0.02* |
| Carbohydrate | 1.04±0.25** | -0.05±0.06 | 0.77±0.28* | -0.09±0.09 | 0.69±0.26* | -0.06±0.10 | 43.05±5.87** | -0.11±0.03* |
| Fat | 0.30±0.05** | 0.30±0.05** | 0.10±0.03** | 0.47±0.07** | 0.11±0.03** | 0.46±0.07** | 24.10±2.15** | 0.04±0.03 |
| **67 ≤ Age ≤ 72 years (n = 634)** | | | | | | | | |
| Protein | 2.37±0.35** | -0.39±0.05** | 2.35±0.47** | -0.53±0.07** | 2.68±0.57** | -0.58±0.08** | 24.73±2.40** | 0.05±0.03 |
| Carbohydrate | 1.08±0.33** | -0.08±0.08 | 0.79±0.32* | -0.10±0.11 | 0.65±0.29* | -0.05±0.11 | 57.33±10.59** | -0.18±0.05** |
| Fat | 0.27±0.05** | 0.31±0.06** | 0.12±0.03** | 0.42±0.08** | 0.13±0.04** | 0.40±0.08** | 18.64±2.24** | 0.12±0.03** |
| **78 ≤ Age ≤ 84 years (n = 497)** | | | | | | | | |
| Protein | 2.68±0.40** | -0.47±0.05** | 3.71±0.82** | -0.77±0.08** | 3.91±0.97** | -0.81±0.09** | 24.11±2.07** | 0.05±0.03 |
| Carbohydrate | 1.20±0.44* | -0.12±0.09 | 0.71±0.40 | -0.12±0.14 | 0.76±0.48 | -0.15±0.16 | 36.64±7.24** | -0.07±0.05 |
| Fat | 0.20±0.04** | 0.39±0.06** | 0.06±0.02* | 0.60±0.10** | 0.04±0.02* | 0.66±0.11** | 26.93±3.08** | 0.004±0.03 |
| **Low BMI (or BMI ≤ 22; n = 110)** | | | | | | | | |
| Protein | 1.29±0.46* | -0.20±0.13 | 1.20±0.63 | -0.34±0.19 | 1.13±0.61 | -0.32±0.20 | 22.24±1.88** | -0.03±0.03 |
| Carbohydrate | 0.53±0.39 | 0.09±0.19 | 0.25±0.28 | 0.16±0.28 | 0.24±0.28 | 0.17±0.29 | 21.33±3.67** | -0.01±0.04 |
| Fat | 0.69±0.31* | 0.02±0.13 | 0.42±0.28 | 0.03±0.19 | 0.43±0.30 | 0.02±0.20 | 18.82±1.98** | 0.02±0.03 |
| **Normal BMI (or 22 < BMI ≤ 27; n = 681)** | | | | | | | | |
| Protein | 2.35±0.31** | -0.40±0.05** | 2.40±0.47** | -0.58±0.07** | 2.71±0.57** | -0.65±0.08** | 23.81±0.71** | 0.02±0.01 |
| Carbohydrate | 0.81±0.26* | -0.01±0.08 | 0.44±0.21* | 0.02±0.12 | 0.37±0.19 | 0.05±0.13 | 27.92±1.86** | -0.03±0.02 |
| Fat | 0.32±0.06** | 0.26±0.05** | 0.14±0.04** | 0.36±0.08** | 0.12±0.04* | 0.39±0.09** | 23.81±0.93** | 0.01±0.01 |
| **High BMI (or BMI> 27; n = 908)** | | | | | | | | |
| Protein | 2.10±0.24** | -0.37±0.04** | 2.53±0.43** | -0.62±0.06** | 2.51±0.42** | -0.59±0.06** | 28.37±1.46** | 0.03±0.02 |
| Carbohydrate | 1.58±0.38** | -0.19±0.06* | 1.15±0.43* | -0.24±0.10* | 1.21±0.44* | -0.23±0.09* | 35.04±3.73** | -0.03±0.03 |
| Fat | 0.19±0.03** | 0.39±0.04** | 0.05±0.01** | 0.61±0.07** | 0.06±0.02** | 0.59±0.07** | 30.48±2.04** | 0.01±0.02 |
| **Lower physical activity quartile (PASE score < 58.57; n = 348)** | | | | | | | | |
| Protein | 2.33±0.39** | -0.42±0.06** | 3.36±0.92** | -0.76±0.10** | 3.07±0.84** | -0.71±0.10** | 29.69±3.68** | -0.01±0.04 |
| Carbohydrate | 0.65±0.25* | -0.03±0.10 | 0.24±0.16 | -0.13±0.17 | 0.29±0.18 | 0.10±0.16 | 35.23±9.18** | -0.05±0.07 |
| Fat | 0.29±0.07** | 0.27±0.06** | 0.10±0.04* | 0.41±0.12** | 0.11±0.04* | 0.40±0.11** | 25.14±4.04** | 0.04±0.05 |
| **Middle physical activity quartiles (58.57 ≤ PASE score ≤ 124.99; n = 877)** | | | | | | | | |
| Protein | 1.90±0.22** | -0.34±0.05** | 1.96±0.31** | -0.49±0.06** | 2.07±0.36** | -0.52±0.06** | 23.23±1.72** | 0.06±0.03* |
| Carbohydrate | 1.20±0.31** | -0.12±0.07 | 0.96±0.33* | -0.16±0.09 | 0.88±0.33* | -0.15±0.10 | 38.74±5.99** | -0.09±0.04* |

| | | | | | | | | |
|---|---|---|---|---|---|---|---|---|
| Fat | 0.27±0.04** | 0.29±0.05** | 0.12±0.03** | 0.42±0.06** | 0.11±0.03** | 0.43±0.07** | 26.00±2.50** | 0.02±0.03 |
| **Higher physical activity quartile (PASE score > 124.99; n = 473)** | | | | | | | | |
| Protein | 2.27±0.36** | -0.36±0.06** | 2.31±0.53** | -0.54±0.08** | 2.57±0.64** | -0.59±0.09** | 24.67±2.33** | 0.04±0.03 |
| Carbohydrate | 1.36±0.48* | -0.13±0.09 | 1.01±0.53 | -0.17±0.13 | 0.74±0.42 | -0.09±0.15 | 59.73±11.79** | -0.20±0.05** |
| Fat | 0.27±0.06** | 0.32±0.06** | 0.10±0.03* | 0.48±0.09** | 0.11±0.04* | 0.45±0.10** | 17.73±2.19** | 0.13±0.04** |
| **Without Diabetes (n = 1512)** | | | | | | | | |
| Protein | 2.10±0.19** | -0.36±0.03** | 2.19±0.26** | -0.52±0.04** | 2.31±0.30** | -0.54±0.05** | 24.72±1.40** | 0.04±0.02* |
| Carbohydrate | 1.31±0.26** | -0.13±0.05* | 0.99±0.27** | -0.16±0.07* | 0.93±0.27** | -0.15±0.07* | 40.28±4.90** | -0.10±0.03* |
| Fat | 0.26±0.03** | 0.32±0.03** | 0.12±0.02** | 0.44±0.05** | 0.11±0.02** | 0.44±0.05** | 24.03±1.75** | 0.04±0.02 |
| **With Diabetes (n = 187)** | | | | | | | | |
| Protein | 2.56±0.67** | -0.43±0.09** | 3.51±1.57** | -0.77±0.16** | 3.21±1.36* | -0.70±0.15** | 33.69±5.34** | -0.05±0.06 |
| Carbohydrate | 0.64±0.34 | -0.04±0.14 | 0.26±0.24 | -0.10±0.24 | 0.32±0.28 | 0.08±0.23 | 30.70±8.96** | -0.01±0.08 |
| Fat | 0.27±0.10* | 0.29±0.11* | 0.07±0.05 | 0.49±0.20* | 0.08±0.06 | 0.46±0.19* | 27.80±6.07** | 0.02±0.06 |
| **Never smoked (n = 883)** | | | | | | | | |
| Protein | 2.03±0.24** | -0.36±0.04** | 2.30±0.42** | -0.60±0.07** | 2.28±0.39** | -0.56±0.06** | 22.07±1.64** | 0.08±0.03* |
| Carbohydrate | 0.89±0.25** | -0.04±0.07 | 0.53±0.24* | -0.05±0.11 | 0.56±0.24* | -0.03±0.11 | 41.02±6.89** | -0.10±0.04* |
| Fat | 0.28±0.05** | 0.28±0.05** | 0.09±0.02** | 0.46±0.08** | 0.11±0.03** | 0.42±0.07** | 26.94±2.73** | 0.01±0.03 |
| **Current smoker (n = 85)** | | | | | | | | |
| Protein | 4.69±1.85* | -0.66±0.14** | 8.62±5.78 | -1.09±0.25** | 8.63±5.83 | -1.09±0.24** | 31.49±6.16** | -0.06±0.07 |
| Carbohydrate | 0.49±0.47 | 0.11±0.24 | 0.30±0.49 | 0.09±0.43 | 0.27±0.45 | 0.12±0.43 | 24.50±10.05* | 0.02±0.11 |
| Fat | 0.15±0.11 | 0.45±0.20* | 0.01±0.01 | 1.08±0.37* | 0.01±0.01 | 1.03±0.38* | 25.42±7.75* | 0.01±0.09 |
| **Former smoker (n = 702)** | | | | | | | | |
| Protein | 2.16±0.27** | -0.36±0.05** | 2.03±0.37** | -0.51±0.06** | 2.39±0.48** | -0.58±0.07** | 27.53±2.30** | 0.01±0.03 |
| Carbohydrate | 1.42±0.37** | -0.15±0.07* | 1.09±0.41* | -0.20±0.10* | 0.90±0.38* | -0.16±0.11 | 47.70±7.87** | -0.14±0.04* |
| Fat | 0.27±0.04** | 0.31±0.05** | 0.11±0.03** | 0.44±0.07** | 0.10±0.03** | 0.44±0.08** | 20.31±2.08** | 0.09±0.03* |

**Table S2**. Summary of mixture experiment models (first Scheffé's model) assessing the linear effects of the EC from each macronutrients on TEI, height-weight adjusted-TEI and BMI. Slope estimates with their standard errors (all coefficients are highly significant, p-value < 0.001)

| | EC from macronutrients (%) | TEI | Adjusted-TEI | BMI |
|---|---|---|---|---|
| **All individuals** | | | | |
| | Protein | -77±12 | -100±11 | 0.34±0.03 |
| | Carbohydrate | 84±4 | 61±3 | 0.25±0.01 |
| | Fat | 141±5 | 113±5 | 0.30±0.01 |
| **Women** | | | | |
| | Protein | -51±14 | -80±13 | 0.32±0.04 |
| | Carbohydrate | 77±4 | 50±4 | 0.25±0.01 |
| | Fat | 118±6 | 91±6 | 0.30±0.02 |
| **Men** | | | | |
| | Protein | -93±17 | -123±16 | 0.36±0.03 |
| | Carbohydrate | 94±5 | 63±5 | 0.24±0.01 |
| | Fat | 156±8 | 126±7 | 0.29±0.02 |
| **67 ≤ Age ≤ 72 years** | | | | |
| | Protein | -93±20 | -108±19 | 0.33±0.05 |
| | Carbohydrate | 89±6 | 63±6 | 0.23±0.02 |
| | Fat | 149±9 | 119±8 | 0.34±0.02 |
| **78 ≤ Age ≤ 84 years** | | | | |
| | Protein | -95±20 | -130±18 | 0.35±0.04 |
| | Carbohydrate | 82±7 | 56±6 | 0.25±0.01 |
| | Fat | 147±9 | 113±8 | 0.27±0.02 |
| **Low BMI (BMI ≤ 22)** | | | | |
| | Protein | -77±25 | -90±29 | 0.16±0.03 |
| | Carbohydrate | 90±15 | 64±7 | 0.21±0.01 |
| | Fat | 88±21 | 99±10 | 0.22±0.01 |
| **Normal BMI (22 < BMI ≤ 27)** | | | | |
| | Protein | -89±19 | -106±17 | 0.27±0.01 |
| | Carbohydrate | 94±6 | 65±6 | 0.24±0.01 |
| | Fat | 138±9 | 101±8 | 0.25±0.01 |
| **High BMI (BMI > 27)** | | | | |
| | Protein | -75±15 | -111±14 | 0.35±0.03 |
| | Carbohydrate | 75±5 | 48±4 | 0.30±0.01 |
| | Fat | 151±7 | 118±6 | 0.31±0.01 |
| **Lower physical activity quartile (PASE score < 58.57)** | | | | |
| | Protein | -76±21 | -111±19 | 0.27±0.06 |
| | Carbohydrate | 86±7 | 58±6 | 0.28±0.02 |
| | Fat | 128±10 | 93±10 | 0.31±0.03 |
| **Middle physical activity quartiles (58.57 ≤ PASE score ≤ 124.99)** | Protein | -61±16 | -86±15 | 0.37±0.04 |
| | Carbohydrate | 81±5 | 58±5 | 0.25±0.01 |
| | Fat | 133±7 | 108±7 | 0.28±0.02 |
| **Higher physical activity quartile (PASE score > 124.99)** | Protein | -92±24 | -114±22 | 0.31±0.05 |
| | Carbohydrate | 89±8 | 60±7 | 0.22±0.01 |
| | Fat | 158±11 | 123±10 | 0.34±0.02 |
| **Without Diabetes** | | | | |
| | Protein | -74±13 | -96±11 | 0.33±0.03 |
| | Carbohydrate | 83±4 | 60±4 | 0.25±0.01 |
| | Fat | 142±6 | 114±5 | 0.29±0.01 |
| **With Diabetes** | | | | |
| | Protein | -86±32 | -114±29 | 0.23±0.08 |
| | Carbohydrate | 87±11 | 53±10 | 0.30±0.03 |
| | Fat | 139±16 | 99±15 | 0.32±0.04 |
| **Never smoked** | | | | |
| | Protein | -76±16 | -107±15 | 0.40±0.04 |
| | Carbohydrate | 85±5 | 56±5 | 0.24±0.01 |
| | Fat | 133±8 | 101±7 | 0.27±0.02 |
| **Current smoker** | | | | |
| | Protein | -162±50 | -195±48 | 0.22±0.09 |

| | | | | |
|---|---|---|---|---|
| | Carbohydrate | 89±18 | 51±18 | 0.28±0.03 |
| | Fat | 169±27 | 144±26 | 0.28±0.05 |
| **Former smoker** | | | | |
| | Protein | -67±18 | -81±16 | 0.28±0.04 |
| | Carbohydrate | 84±6 | 54±5 | 0.24±0.01 |
| | Fat | 145±7 | 107±7 | 0.33±0.02 |

**Table S3.** Linear effects of EC from protein and from carbohydrate:fat (CF; transformed via PCA) on TEI, adjusted-TEI and BMI. Ordinary least squares estimates of parameters with their standard errors (se) andp-values (** for p ≤ 0.001 and * for 0.001 < p ≤ 0.05)

| | EC from macronutrients | TEI | Adjusted-TEI | BMI |
|---|---|---|---|---|
| **All individuals** | | | | |
| | Protein | -189±14** | -187±13** | 0.07±0.03* |
| | PCA-transformed CF | -258±35** | -234±32** | -0.24±0.08* |
| **Women** | | | | |
| | Protein | -148±16** | -151±16** | 0.04±0.05 |
| | PCA-transformed CF | -184±40** | -184±40 | -0.23±0.12 |
| **Men** | | | | |
| | Protein | -218±20** | -217±19** | 0.09±0.04* |
| | PCA-transformed CF | -284±51** | -289±49** | -0.23±0.10* |
| **67 ≤ Age ≤ 72 years** | | | | |
| | Protein | -211±24** | -198±23** | 0.04±0.06 |
| | PCA-transformed CF | -271±58** | -250±54** | -0.50±0.14** |
| **78 ≤ Age ≤ 84 years** | | | | |
| | Protein | -210±23** | -215±21** | 0.09±0.05 |
| | PCA-transformed CF | -296±63** | -261±57** | -0.11±0.14 |
| **Low BMI (BMI ≤ 22)** | | | | |
| | Protein | -188±30** | -182±27** | -0.05±0.04 |
| | PCA-transformed CF | -178±71* | -136±65* | -0.06±0.09 |
| **Normal BMI (22 < BMI ≤ 27)** | | | | |
| | Protein | -205±23** | -189±21** | 0.02±0.02 |
| | PCA-transformed CF | -191±56** | -159±51** | -0.05±0.04 |
| **High BMI (BMI > 27)** | | | | |
| | Protein | -187±18** | -194±17** | 0.05±0.03 |
| | PCA-transformed CF | -350±46** | -322±42** | -0.05±0.09 |
| **Lower physical activity quartile (PASE score < 58.57)** | | | | |
| | Protein | -182±25** | -186±23** | -0.03±0.07 |
| | PCA-transformed CF | -198±69* | -163±64* | -0.18±0.20 |
| **Middle physical activity quartiles (58.57 ≤ PASE score ≤ 124.99)** | Protein | -168±19** | -168±18** | 0.11±0.04* |
| | PCA-transformed CF | -236±47** | -229±43** | -0.14±0.11 |
| **Higher physical activity quartile (PASE score > 124.99)** | | | | |
| | Protein | -215±29** | -294±27** | 0.03±0.06 |
| | PCA-transformed CF | -307±70** | -282±64** | -0.53±0.13** |
| **Without Diabetes** | | | | |
| | Protein | -186±15** | -183±14** | 0.06±0.03 |
| | PCA-transformed CF | -262±37** | -241±34** | -0.21±0.08* |
| **With Diabetes** | | | | |
| | Protein | -198±39** | -189±36** | -0.08±0.09 |
| | PCA-transformed CF | -231±103* | -205±96* | -0.10±0.25 |
| **Never smoked** | | | | |
| | Protein | -185±19** | -185±18** | 0.14±0.05* |
| | PCA-transformed CF | -208±48** | -195±45** | -0.13±0.11 |
| **Current smoker** | | | | |
| | Protein | -288±60** | -290±58** | -0.06±0.11 |
| | PCA-transformed CF | -362±178* | -418±174* | -0.01±0.32 |
| **Former smoker** | | | | |
| | Protein | -181±21** | -161±19** | -0.01±0.05 |
| | PCA-transformed CF | -285±52** | -250±47** | -0.41±0.12** |

**Table S4**. Relationship between BMI (Kg.m$^{-2}$) and unadjusted TEI (kJ) over time. Ordinary least squares (OLS) estimates with standard errors (and correlations). All OLS estimates are multiplied by $10^{-5}$, and (*) for significant estimates.

| | All individuals | 67 ≤ Age < 72 years | 78 ≤ Age ≤ 84 years |
|---|---|---|---|
| **For $i = 1$** | | | |
| $TEI_i$ vs $\Delta_{BMI_{[i+1,i]}}$ | 4.24±8.15 (-0.014) | 14.82±13.41 (0.048) | -9.29±16.61 (-0.028) |
| $TEI_i$ vs $\Delta_{BMI_{[4,i]}}$ | -0.84±8.71 (-0.003) | 20.47±14.17 (0.064) | -0.13±16.67 (-0.000) |
| $\Delta_{TEI_{[i+1,i]}}$ vs $\Delta_{BMI_{[i+1,i]}}$ | -5.58±5.55 (-0.027) | -5.36±8.57 (-0.027) | -3.03±11.41 (-0.014) |
| $\Delta_{TEI_{[4,i]}}$ vs $\Delta_{BMI_{[4,i]}}$ | -4.10±5.75 (-0.020) | -4.86±9.29 (-0.023) | -7.12±10.34 (-0.037) |
| **For $i = 2$** | | | |
| $TEI_i$ vs $\Delta_{BMI_{[i+1,i]}}$ | -3.45±7.76 (0.012) | -13.97±12.96 (-0.047) | 12.69±13.96 (0.048) |
| $TEI_i$ vs $\Delta_{BMI_{[4,i]}}$ | 8.69±8.17 (0.029) | 11.54±13.42 (0.038) | 3.63±15.11 (0.013) |
| $\Delta_{TEI_{[i+1,i]}}$ vs $\Delta_{BMI_{[i+1,i]}}$ | -7.69±5.72 (-0.036) | 3.35±9.01 (0.016) | -15.92±11.24 (-0.074) |
| $\Delta_{TEI_{[4,i]}}$ vs $\Delta_{BMI_{[4,i]}}$ | -11.56±5.68* (-0.056) | -17.01±9.36 (-0.081) | -9.05±10.51 (-0.046) |
| **For $i = 3$** | | | |
| $TEI_i$ vs $\Delta_{BMI_{[i+1,i]}}$ | 4.73±8.48 (-0.015) | -3.02±9.73 (-0.062) | 7.93±15.54 (0.027) |
| $\Delta_{TEI_{[i+1,i]}}$ vs $\Delta_{BMI_{[i+1,i]}}$ | -10.46±5.72 (-0.051) | -4.91±14.07 (-0.014) | 17.18±10.47 (-0.087) |

$i$ indicates annual visit (1 to 4: recruitment to third year of follow-up).

**Table S5.** AIC values for the five Scheffé's mixture experiment models on unadjusted TEI

| | Model 1 | Model 2 | Model 3 | Model 4 | Model 5 |
|---|---|---|---|---|---|
| **All individuals** | 30 758.02 | 30 549.90 | 30 554.75 | 30 554.33 | 30 556.70 |
| **Women** | 15 701.74 | 15 610.77 | 15 616.34 | 15 618.36 | 15 618.33 |
| **Men** | 14 777.40 | 14 655.24 | 14 659.99 | 14 665.55 | 14 661.50 |
| **67 ≤ Age ≥ 72 years** | 11 491.01 | 11 411.50 | 11 414.44 | 11 419.97 | 11 416.21 |
| **78 ≤ Age ≥ 84 years** | 8 953.20 | 8 866.26 | 8 869.53 | 8 876.42 | 8 870.53 |
| **Low BMI (BMI ≤ 22)** | 1 984.49 | 1 982.82 | 1 990.21 | 1 990.96 | 1 992.20 |
| **Normal BMI (22 < BMI ≤ 27)** | 12 360.40 | 12 279.14 | 12 284.25 | 12 285.38 | 12 283.24 |
| **High BMI (BMI > 27)** | 16 413.34 | 16 277.42 | 16 281.88 | 16 282.47 | 16 283.28 |
| **Lower physical activity quartile (PASE score < 58.57)** | 6 228.01 | 6 178.76 | 6 182.85 | 6 187.65 | 6 182.80 |
| **Middle physical activity quartiles (58.57 ≤ PASE score ≥ 124.99)** | 15 812.83 | 15 724.67 | 15 729.22 | 15 730.78 | 15 730.56 |
| **Higher physical activity quartile (PASE score > 124.99)** | 8 628.64 | 8 568.89 | 8 573.94 | 8 577.89 | 8 575.20 |
| **Without Diabetes** | 27 380.78 | 27 201.12 | 27 205.75 | 27 204.36 | 27 207.26 |
| **With Diabetes** | 3 378.09 | 3 355.53 | 3 361.24 | 3 366.53 | 3 362.89 |
| **Never smoker** | 15 980.06 | 15 885.43 | 15 889.38 | 15 891.44 | 15 890.71 |
| **Current smoker** | 1 570.04 | 1 551.84 | 1 553.61 | 1 557.55 | 1 554.83 |
| **Former smoker** | 12 674.53 | 12 587.34 | 12 591.45 | 12 595.98 | 12 593.44 |

The five Scheffé's mixture experiment models are defined below.

### Overview on mixture experiments models

(28) defined four mixture models and two mixture process models to perform linear regression with correlated predictors (27). In this section, we presented the five models used in this paper including the four mixture models.

The null linear model is a linear regression with only the intercept (EqS.1),

$$TEI = 1 + \varepsilon, \tag{EqS.1}$$

where $\varepsilon$ is the model residuals.

The first Scheffé's model is a linear model (EqS.2), of which coefficients, in our case, are the expected total energy intake (TEI) when the three ECs are unitary. When ECs are greater than 1, they represent the decrease or increase value of TEI for every unitary increase of ECs.

$$TEI = \sum_{i=1}^{q} \beta_i EC_i + \varepsilon, \tag{EqS.2}$$

where $EC_i$ is the energy contribution of the *i*th macronutrient (i = 1 … *q*; *q* = 3) and $\beta_i$ is the effect of $EC_i$ on TEI.

The quadratic model (second Scheffé's model) adds a part of quadratic effects of pairs of contributions ($\gamma_{ij}$), which represent the increase or decrease of the instantaneous slope between TEI and ECs, for a unitary increase of ECs (EqS.3).

$$TEI = \sum_{i=1}^{q} \beta_i EC_i + \sum_{i=1}^{q-1} \sum_{j=i+1}^{q} \gamma_{ij} EC_i EC_j + \varepsilon, \tag{EqS.3}$$

The full cubic (EqS.4) (third Scheffé's model) and the special-cubic models (fourth Scheffé's model) are popular alternatives to the quadratic model.

$$TEI = \sum_{i=1}^{q} \beta_i EC_i + \sum_{i=1}^{q-1} \sum_{j=i+1}^{q} \gamma_{ij} EC_i EC_j + \sum_{i=1}^{q-1} \sum_{j=i+1}^{q} \delta_{ij} EC_i EC_j (EC_i - EC_j) +$$
$$\sum_{i=1}^{q-2} \sum_{j=i+1}^{q-1} \sum_{k=j-1}^{q} \gamma_{ijk} EC_i EC_j EC_k + \varepsilon, \tag{EqS.4}$$

The special-cubic model is the full cubic model without the $\delta_{ij}EC_iEC_j(EC_i - EC_j)$ term (EqS.5).

$$TEI = \sum_{i=1}^{q} \beta_i EC_i + \sum_{i=1}^{q-1} \sum_{j=i+1}^{q} \gamma_{ij} EC_i EC_j + \sum_{i=1}^{q-2} \sum_{j=i+1}^{q-1} \sum_{k=j-1}^{q} \gamma_{ijk} EC_i EC_j EC_k + \varepsilon$$

(EqS.5)
